# FOXP3 mRNA profile prognostic of T cell mediated rejection and human kidney allograft survival

**DOI:** 10.1101/2020.04.28.20083493

**Authors:** Danny Luan, Darshana M. Dadhania, Ruchuang Ding, Thangamani Muthukumar, Michelle Lubetzky, John R. Lee, Vijay K. Sharma, Phyllis August, Franco B. Mueller, Joseph E. Schwartz, Manikkam Suthanthiran

## Abstract

**Background and objectives:** T cell mediated rejection (TCMR) is the most frequent type of acute rejection and is associated with kidney allograft failure. Almost 40% of TCMR episodes fail to respond to anti-rejection therapy. FOXP3 is a specification factor for regulatory T cells and our single center study of 83 kidney allograft recipients showed that urinary cell FOXP3 mRNA level is diagnostic of TCMR and predicts TCMR reversibility and allograft survival. The objective of the current study is to determine whether our original findings could be replicated in an independent cohort of 480 kidney allograft recipients enrolled in the multicenter Clinical Trials of Organ Transplantation (CTOT)-04.

**Design, setting, participants, and measurements:** We measured levels of FOXP3 mRNA and levels of mRNA for CD25, CD3E, and perforin, and 18S rRNA in 3559 urines from 480 kidney allograft recipients prospectively enrolled in CTOT-04. RNA was isolated from the urinary cells and preamplification-enhanced real-time quantitative PCR assays were used to measure mRNAs.

**Results:** 18S rRNA normalized levels of mRNA for FOXP3 (*P*=0.01, Kruskal-Wallis test), CD25 (*P*=0.01), CD3E (*P*<0.0001), and perforin (*P*<0.0001) distinguished patients with TCMR biopsies from those with No Rejection biopsies and those with stable graft function. FOXP3 mRNA level, but not the levels of mRNA for CD25, CD3E, or perforin, predicted TCMR reversal (AUC=0.764; 95% confidence interval, 0.611 to 0.917; *P*=0.008). Multivariable logistic regression analysis showed that FOXP3 mRNA level remains predictive after adjustment for potential cofounders. Kaplan-Meier survival curve analysis showed that FOXP3 mRNA level (*P*=0.0306) but not the levels of mRNA for CD25, CD3E, or perforin, is associated with kidney allograft survival.

**Conclusion:** In the independent CTOT-04 cohort, we demonstrate that urinary cell level of FOXP3 mRNA is diagnostic of TCMR, predicts its reversibility, and is prognostic of kidney allograft survival following an episode of TCMR.

## Introduction

T cell mediated rejection (TCMR) is the most frequent type of acute rejection (*1–4*). Anti-rejection therapy has evolved over the years, but Bouatou *et al* found in a comprehensive study of 256 kidney allograft recipients with kidney allograft biopsies showing TCMR that 40% of TCMR episodes fail to respond to anti-rejection treatment (*5*). Moreover, the histologic hallmarks of TCMR – Banff tubulitis score and the interstitial inflammation score at the time of TCMR biopsy diagnosis– were not determinants of kidney allograft loss whereas the inflammation score and peritubular capillaritis score observed in the biopsy performed 3 months after anti-rejection treatment were independent predictors of kidney allograft failure (*5*). The need for post-treatment parameters to improve prognostic accuracy is problematic from the perspective of clinical decision making at the time of TCMR diagnosis. Also, the need for multiple biopsies – one to diagnose TCMR and a second one to better prognosticate TCMR outcome – is challenging in view of complications related to the invasive biopsy procedure; the well-documented inter-observer variability in the grading of biopsies is yet another problem *(6, 7)*. Because molecular or cellular mechanisms for recalcitrant TCMR have not been deciphered, a mechanistic approach to treatment of TCMR is currently lacking, and often the “one-size-fits all” strategy is followed in the clinical setting.

To fill the existing gaps and improve our understanding of mechanisms for non-responsiveness to anti-rejection treatment, we investigated whether urinary cell FOXP3 mRNA profiles predict TCMR reversal. Our approach was informed by the following considerations: (i) our earlier single center study of 83 kidney allograft recipients demonstrating that urinary cell level of FOXP3 mRNA predicts TCMR reversibility and identifies patients at risk for kidney allograft failure (*8*); (ii) naturally occurring CD4+FOXP3+ regulatory T cells (Tregs) play a non-redundant role in immune homeostasis and in the maintenance of self-tolerance (*9, 10*); (iii) preclinical data showing that Tregs prevent rejection and promote transplant tolerance (*11–13*); and (iv) consideration of adoptive Treg therapy in the clinic to control autoimmunity or promote tolerance to allografts (*14, 15*). We also considered that it is important to investigate the reproducibility of the associations we previously reported for urinary cell FOXP3 mRNA levels (*8*) in view of the reproducibility crisis in the replication of scientific findings (*16, 17*).

In the current investigation, we measured levels of mRNAs in 3505 urine specimens from 480 kidney transplant recipients enrolled in the multicenter Clinical Trials in Organ Transplantation 04 (CTOT-04) study in order to determine whether urinary cell FOXP3 gene expression profile is diagnostic of TCMR, predicts TCMR reversibility, and is prognostic of kidney allograft survival following an episode of TCMR.

## Materials and Methods

### Study subjects

In the parent CTOT-04 study, 485 kidney allograft recipients were prospectively enrolled at 5 transplant sites. Urine was prospectively collected on days 3, 7, 15, and 30 and in months 2, 3, 4, 5, 6, 9 and 12 as well as the time of each kidney allograft biopsy and 2 weeks thereafter. Urine cell pellets were prepared at each clinical site and shipped to the Gene Expression Monitoring (GEM) core at Weill Cornell Medicine. The GEM core isolated RNA from the urinary cell pellets, reverse transcribed it to cDNA, and checked for transcript quality thresholds – 100 copies of TGFB1 mRNA and 5×10^7^ copies of 18S rRNA per one microgram of total RNA – prior to downstream data analysis. Absolute levels of mRNA for CD3E, perforin, granzyme B, proteinase inhibitor-9 (PI-9), CD103, interferon inducible protein-10 (IP-10), CXCR3, TGFB1, and 18S rRNA (CTOT-04 Prespecified mRNA Panel) were measured. A total of 3559 urine specimens from 485 kidney allograft recipients passed the transcript quality thresholds. The primary objectives of the parent CTOT-04 study were to determine whether urinary cell levels of a prespecified mRNA panel, measured at time of biopsy, are diagnostic of TCMR and whether the levels in sequential samples predict future development of TCMR. The parent CTOT-04 study did not investigate whether the measured transcripts predict TCMR reversal or kidney allograft survival. Urinary cell levels of FOXP3 mRNA and mRNA for CD25 were not measured in the parent CTOT-04 study.

In the current study, cDNA prepared from the same 3559 urine cell pellets were retrieved for the measurement of FOXP3 mRNA and CD25 mRNA. Prior to measurement of mRNAs, the cDNA was again assessed for transcript quality thresholds and 3505 of the 3559 cDNAs (98.5%) prepared from the urine specimens from 480 of 485 kidney allograft recipients from the parent CTOT-04 study cohort met the quality threshold, and were used to measure urinary cell mRNA levels using preamplification-enhanced real-time quantitative polymerase chain reaction (customized RT-QPCR) assays developed in our laboratory (*4*). Table S1 lists the oligonucleotide primers and TaqMan probes we designed for the absolute quantification of mRNAs. Additional details of the customized RT-QPCR assay developed in our laboratory and used to measure transcript levels have been reported (*4*).

In the current investigation, we applied the same criterion we used in our earlier study (*8*) to classify an episode of TCMR as reversible or nonreversible. Specifically, a TCMR episode was classified as reversible if the serum creatinine level returned to within 15 percent of the pre-rejection level within 4 weeks after initiation of anti-rejection therapy.

### Statistical methods

Copy numbers for each mRNA were analyzed after normalization with 18S rRNA copies (x10^−6^) and log_10_-transformation. Kruskal-Wallis and Mann-Whitney statistical tests were used to compare 18S rRNA normalized mRNA levels across diagnoses. Receiver-operating-characteristic (ROC) curves were used to determine the predictive accuracy of urinary cell mRNA levels and the optimal sensitivity and specificity combination was determined based on Youden’s index (*18*). Multivariable logistic regression was used to evaluate urinary cell levels of mRNA after controlling for potential confounders.

Kaplan-Meier survival curves were used to evaluate graft survival rates and to determine the ability of urinary cell mRNA level, measured at the time of biopsy, to anticipate graft failure. Time to event was calculated from time of TCMR biopsy until graft failure or censoring. Subjects were censored if they died prior to experiencing graft failure or were lost to follow-up. Log-rank tests were used to compare survival curves across strata. Long-term follow-up information on graft outcomes, beyond the 3-year duration of the CTOT-04 study, was obtained from the Organ Procurement and Transplantation Network (OPTN). Multivariable Cox proportional hazards regression was used to evaluate urinary cell mRNA as a predictor of graft failure after controlling for potential confounders.

All analyses were performed with SAS software, version 9.4 (SAS Institute) except for the creation of ROC curves using the *pROC* R package (*19*). The datasets and code generated during and/or analyzed during the current study are available from the corresponding author on reasonable request.

### Study approval

The institutional review board at Weill Cornell Medicine approved the study, “Multicenter Study: Use of PCR to Evaluate Immune Regulatory Molecules”, Protocol Number 9608002317.

## Results

### Study groups

Figure 1 shows the distribution of 3505 urine samples collected from the 480 patients and with urinary cell mRNA data. Among the CTOT-04 cohort of 480 kidney allograft recipients, 218 underwent kidney allograft biopsies, which were classified by the on-site pathologist using the Banff classification schema (*20*). Among 262 patients who did not undergo a biopsy, 199 patients contributed 1524 urine samples and met the criteria for stable graft function based on average study recorded serum creatinine values ≤ 2.0 mg/dl, no treatment for acute rejection, and no evidence of cytomegalovirus or polyomavirus type BK infection during the first 12 months post-transplantation (Stable).

**Fig 1.**
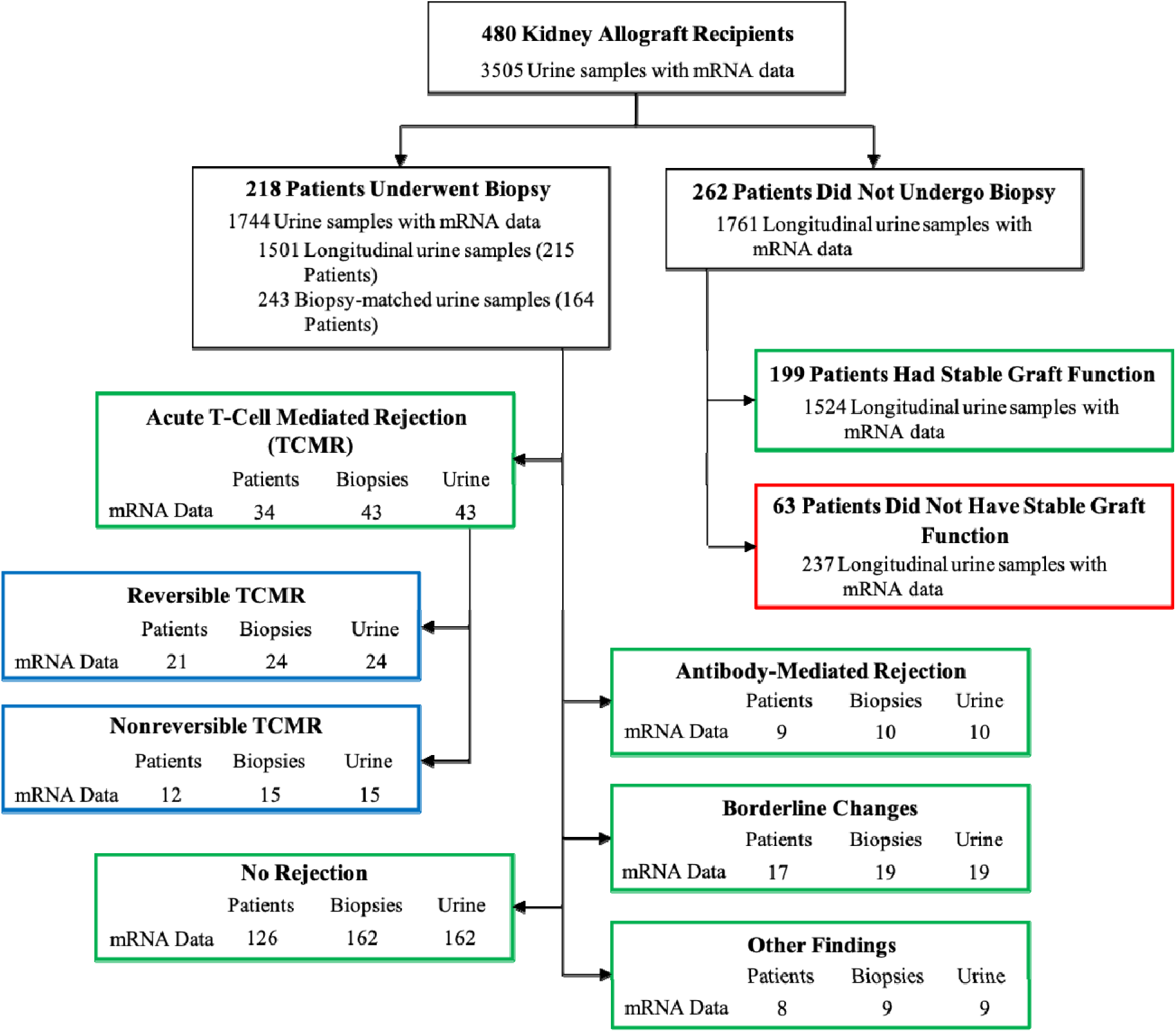
Patients, biopsy results, and urine samples. The distribution of 3505 urine samples from m 480 kidney allograft recipients enrolled in the CTOT-04 study is shown. The number of patients with biopsy-matched urine samples (urine collected from 3 days before to 1 day after biopsy) are shown for patients with TCMR (defined as Banff grade 1A or higher), antibody-mediated rejection, borderline changes, other findings, and those without any rejection features in the biopsy (No Rejection). Among those with TCMR biopsies, 39 of the 43 episodes of TCMR were further classified into those who successfully responded to anti-rejection therapy (Reversible TCMR) and those who failed to respond to anti-rejection therapy (Nonreversible TCMR). A TCMR episode was reversible if serum creatinine returned to within 15% of pre-rejection level within 4 weeks after initiation of anti-rejection therapy. Three patients contributed more than one TCMR biopsy and each contributed ≥1 reversible and ≥1 nonreversible biopsy. Each patient is classified by the status of their last TCMR biopsy. Among patients who did not undergo a biopsy, 199 patients met criteria for stable graft function (listed in Materials and Methods) and contributed 1524 urine samples. Sixty-three patients failed to meet criteria for stable graft function and contributed 237 urine specimens. Green and blue boxes represent samples included in this study, whereas the red box represents samples not included in the data analysis.

### Urinary cell mRNAs diagnostic of TCMR

We compared urinary cell levels of mRNA in 43 urine samples matched to TCMR biopsies from 34 patients, 162 samples matched to No Rejection biopsies from 126 patients, and 1524 samples collected longitudinally from 199 Stable patients. Table 1 is a summary of baseline characteristics of the kidney allograft recipients included in this analysis.

**Table 1.** Characteristics of the kidney allograft recipients and their organ donors.^A^

| Recipient Characteristics | Acute T-Cell Mediated Rejection<br>(n = 34) | No Rejection<br>(n = 126) | Stable<br>(n = 199) |
| --- | --- | --- | --- |
| Biopsy samples | 43 | 162 |  |
| Urine samples | 43 | 162 | 1524 |
| Age, years |  |  |  |
| Mean (SD) | 45.0 (11.8) | 48.0 (13.0) | 49.0 (13.9) |
| Median | 43 | 48 | 50 |
| Min, Max | 24, 73 | <1, 76 | <1, 78 |
| Gender, N (%) |  |  |  |
| Female | 8 (23.5) | 43 (34.1) | 92 (45.8) |
| Male | 26 (76.5) | 83 (65.9) | 109 (54.2) |
| Ethnicity, N (%) |  |  |  |
| Hispanic or Latino | 3 (8.8) | 20 (15.9) | 31 (15.4) |
| Not Hispanic or Latino | 30 (88.2) | 101 (80.2) | 161 (80.1) |
| Unknown or Not Reported | 1 (2.9) | 5 (4.0) | 9 (4.5) |
| Race, N (%) |  |  |  |
| Black or African American | 13 (38.2) | 49 (38.9) | 45 (22.4) |
| White | 20 (58.8) | 62 (49.2) | 119 (59.2) |
| Asian | 1 (2.9) | 9 (7.1) | 10 (5.0) |
| American Indian or Alaska Native | 0 (0) | 0 (0) | 2 (1.0) |
| Other | 0 (0) | 4 (3.2) | 20 (10.0) |
| Unknown or Not Reported | 0 (0) | 2 (1.6) | 5 (2.5) |
| Induction Therapy, N (%) |  |  |  |
| IL-2 Receptor Antibody | 6 (17.6) | 12 (9.5) | 20 (10.1) |
| CAMPATH-1H | 10 (29.4) | 58 (46.0) | 29 (14.6) |
| Thymoglobulin | 15 (44.1) | 38 (30.2) | 135 (67.8) |
| More than one induction therapy | 2 (5.9) | 7 (5.6) | 6 (3.0) |
| No Induction Therapy | 1 (2.9) | 2 (1.6) | 1 (0.5) |
| Missing Information | 0 (0) | 9 (7.1) | 10 (5.0) |
| BMI |  |  |  |
| Mean (SD) | 30.5 (6.2) | 28.5 (6.3) | 27.0 (5.6) |
| Median | 29 | 28 | 26 |
| Min, Max | 22, 43 | 17, 45 | 16, 43 |
| < 18.5 | 0 (0) | 2 (1.6) | 4 (2.0) |
| 18.5 - 24.9 | 9 (26.5) | 35 (27.8) | 58 (28.9) |
| 25.0 - 29.9 | 7 (20.6) | 41 (32.5) | 56 (27.9) |
| ≥ 30.0 | 14 (41.2) | 40 (31.7) | 44 (21.9) |
| Missing | 4 (11.8) | 8 (6.3) | 39 (19.6) |

| Donor Characteristics |  |  |  |
| --- | --- | --- | --- |
| Age, years |  |  |  |
| Mean (SD) | 41.8 (11.5) | 39.8 (14.8) | 38.9 (14.7) |
| Median | 40 | 41 | 39 |
| Min, Max<sup>B</sup> | 20, 65 | 5, 66 | 1, 73 |
| Missing | 0 | 0 | 3 |
| Gender, N (%) |  |  |  |
| Female | 17 (50.0) | 61 (48.4) | 83 (41.3) |
| Male | 17 (50.0) | 65 (51.6) | 118 (58.7) |
| Ethnicity, N (%) |  |  |  |
| Hispanic or Latino | 4 (11.8) | 21 (16.7) | 36 (17.9) |
| Not Hispanic or Latino | 29 (85.3) | 92 (73.0) | 140 (69.7) |
| Unknown or Not Reported | 1 (2.9) | 13 (10.3) | 25 (12.4) |
| Race, N (%) |  |  |  |
| Black or African American | 10 (29.4) | 34 (27.0) | 19 (9.5) |
| White | 22 (64.7) | 82 (65.1) | 154 (76.6) |
| Asian | 1 (2.9) | 3 (2.4) | 5 (2.5) |
| American Indian or Alaska Native | 0 | 1 (0.8) | 1 (0.5) |
| Other | 0 | 0 (0) | 3 (1.5) |
| Unknown or Not Reported | 1 (2.9) | 6 (4.8) | 19 (9.5) |
| Source of Donor, N (%) |  |  |  |
| Deceased | 15 (44.1) | 57 (45.2) | 85 (42.3) |
| Living/related | 9 (26.5) | 39 (31.0) | 73 (36.3) |
| Living/unrelated | 10 (29.4) | 30 (23.8) | 43 (21.4) |
| Cause of Death, N (%) |  |  |  |
| Anoxia | 4 (26.7) | 9 (15.8) | 15 (17.6) |
| Cerebrovascular Accident/Injury/Stroke | 6 (40.0) | 27 (47.4) | 16 (18.8) |
| Head Injury/Trauma | 2 (13.3) | 8 (14.0) | 24 (28.2) |
| Intracranial Bleed | 1 (6.7) | 2 (3.5) | 6 (7.1) |
| Motor Vehicle Accident | 0 (0) | 1 (1.8) | 2 (2.4) |
| Other | 1 (6.7) | 4 (7.0) | 11 (12.9) |
| Unknown | 0 (0) | 3 (5.3) | 10 (11.8) |
| Missing | 1 (6.7) | 3 (5.3) | 1 (1.2) |

<sup>A</sup> Diagnostic categories of Acute T-Cell Mediated Rejection, No Rejection, and Stable are presented. 34 patients underwent 43 kidney allograft biopsies and contributed 43 acute T-cell mediated rejection biopsy-matched urine samples. 126 patients underwent 162 biopsies and contributed 162 No Rejection biopsy-matched urine samples. 199 patients did not undergo a biopsy and contributed 1524 urine samples. These patients were classified as Stable on meeting the following criteria: stable graft function based on recorded serum creatinine values $\leq 2.0$ mg/dl, no treatment for acute rejection, and no evidence of cytomegalovirus or polyomavirus type BK infection during the first 12 months post-transplantation.

Violin plots with in-laid box-and-whisker plots in Figure 2 illustrate the distribution of log_10_-transformed ratios of mRNA to 18S rRNA copies (x10^−6^) per microgram of total RNA. We replicated our earlier findings that FOXP3 mRNA levels and the levels of mRNA for CD25, CD3E, and perforin are all diagnostic of TCMR; the levels of all 4 mRNAs were significantly higher in urine samples matched to TCMR biopsies than in urine samples matched to No Rejection biopsies. The levels of these mRNAs in urine matched to TCMR biopsies were also higher than in urine samples collected longitudinally from Stable patients. Table S2 shows the median and lower and upper quartiles of the log_10_-transformed 18S rRNA normalized ratios of mRNAs for the diagnostic groups of TCMR, No Rejection, Stable and other biopsy diagnoses.

**Fig 2.**
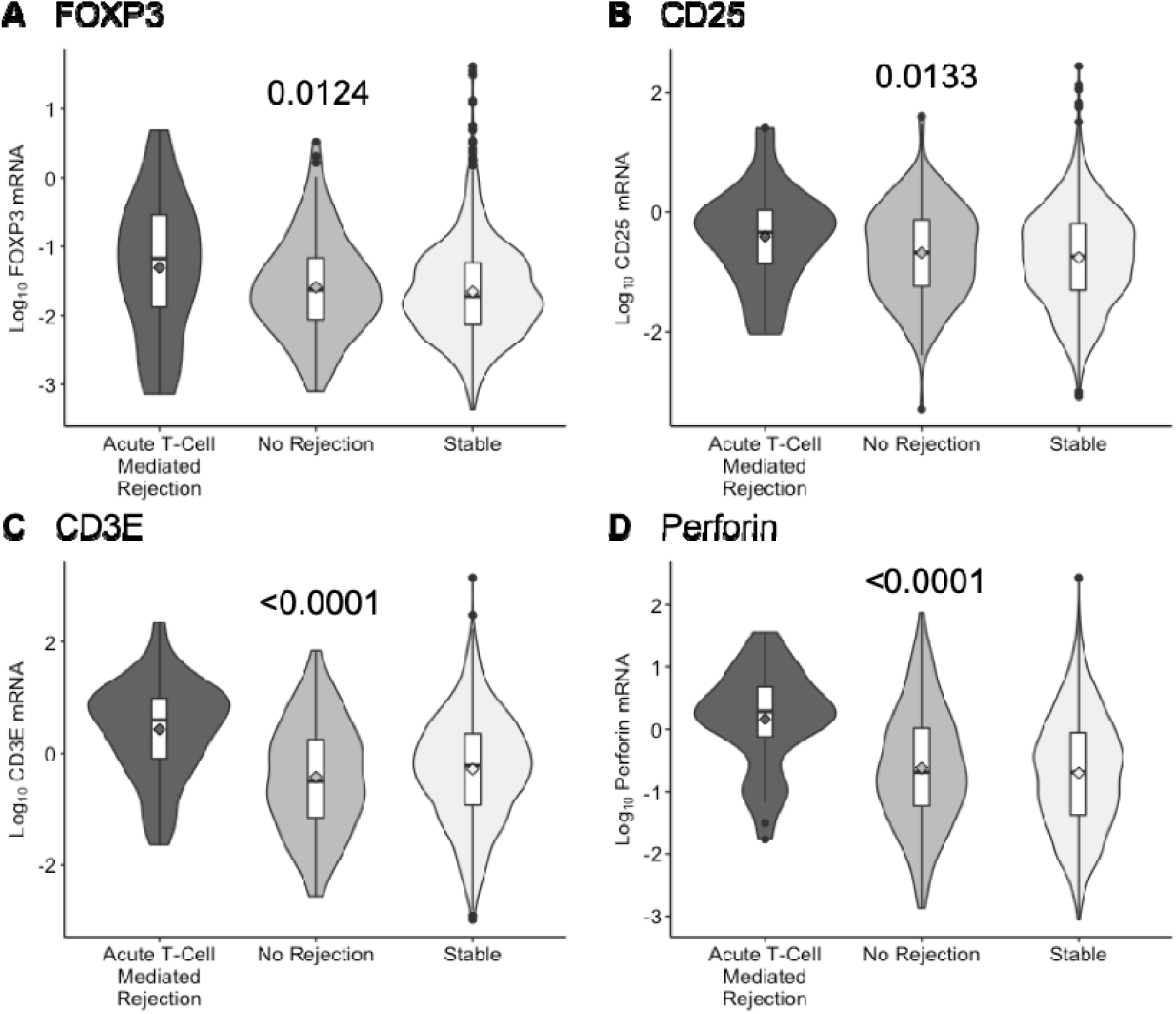
Levels of mRNA in urinary cells. Violin plots with in-laid box-and-whisker plots show the distribution of log_10_-transformed ratios of mRNA copies per microgram of total RNA to 18S ribosomal RNA (rRNA) copies (x10^−6^) per microgram of total RNA for FOXP3, CD25, CD3E, and perforin in 43 urine samples matched to 43 biopsy specimens (from 34 subjects) demonstrating TCMR, 162 urine samples matched to 162 biopsy specimens (from 126 subjects) without any rejection features in the biopsy (No Rejection), and 1524 urine samples collected longitudinally from 199 subjects with stable graft function who did not undergo biopsy (Stable). The in-laid box-and-whisker plots display the 25^th^, 50^th^, and 75^th^ percentile values via the bottom, middle, and top lines in the box, respectively, and the 10^th^ and 90^th^ percentile values via the ends of the bottom and top whiskers, respectively; the diamonds represent the mean and circles indicate outliers. The violin plots display the distribution and spread of observations in each category. The *P*-value from the Kruskal-Wallis test of the null hypothesis of no group differences in the distributions is presented above each set of violin plots.

### Urinary cell mRNA levels stratified by reversibility of TCMR

Among 43 biopsy-confirmed TCMR episodes from 34 patients with biopsy-matched urine samples, 39 episodes from 33 patients were classifiable as reversible (n=24) or nonreversible (n=15); the remaining 4 were not classified due to missing creatinine values (*n*=2), close proximity to BKV nephropathy diagnosis (*n*=1), or close proximity to an earlier biopsy (*n*=1). Tables S2 and S3 list recipient and donor characteristics and biopsy-associated variables, respectively, by TCMR reversible status.

We compared urinary cell levels of mRNAs in urine matched to reversible TCMR biopsies to those matched to nonreversible TCMR biopsies (Table 2). Urinary cell levels of mRNA for FOXP3 (*P*=0.0096, Mann-Whitney Test), but not the levels of mRNA for CD25 (*P*=0.1531), CD3E (*P*=0.1887), or perforin (*P*=0.4322), were significantly higher in urine matched to reversible TCMR biopsies than in samples matched to nonreversible TCMR biopsies.

**Table 2.**
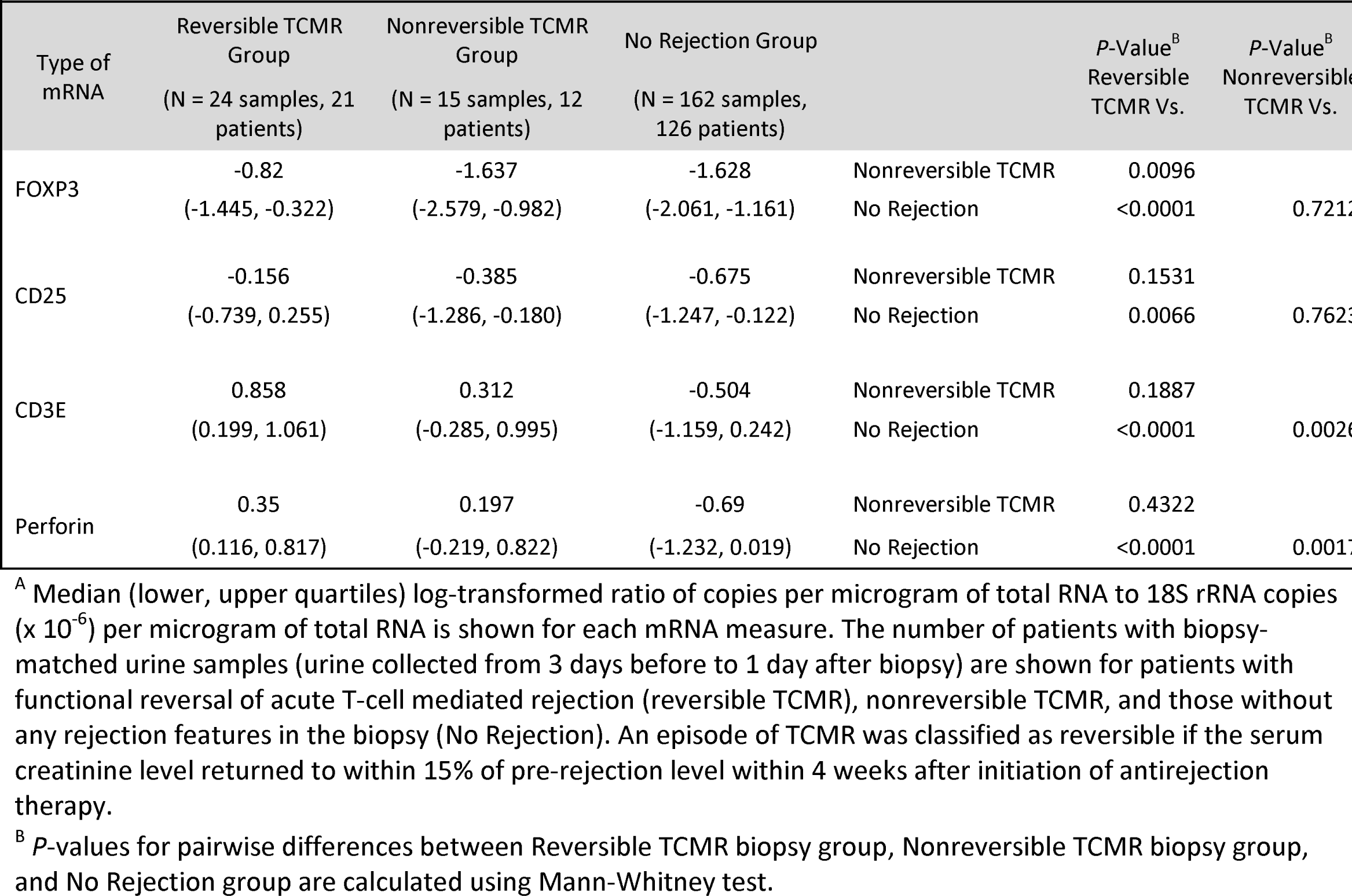
18S rRNA normalized, log_10_-transformed levels of mRNA in urinary cells across reversible and nonreversible TCMR categories.^A^

We also compared levels of mRNAs in urine matched to reversible or nonreversible TCMR biopsies to levels in urine matched to No Rejection biopsies (Table 2). These comparisons identified additional differential gene expression patterns. Urinary cell levels of mRNA for Treg specification factor FOXP3 (*P*<0.0001, Mann-Whitney Test) and CD25 (*P*=0.0066) were significantly higher in urine matched to reversible TCMR biopsies than in urine matched to No Rejection biopsies. In contrast, levels of FOXP3 mRNA (*P*=0.7212) and CD25 mRNA (*P*=0.7623) were not different between urine matched to nonreversible TCMR biopsies and urine matched to No Rejection biopsies. The levels of mRNA encoding cytotoxic perforin and pan T cell specific CD3E were significantly higher both in urine matched to reversible and nonreversible TCMR biopsies compared to urine matched to No Rejection biopsies (Table 2).

### ROC curve analysis of reversal of TCMR

ROC curve analysis yielded an AUC of 0.764 (95% confidence interval [CI], 0.611 to 0.917; *P*=0.008) for the 18S rRNA normalized values of FOXP3 mRNA (Figure 3A). The cut-point that maximized Youden’s index (*18*) was 0.06, yielding a sensitivity of 92% (95% CI, 73% to 99%) and specificity of 87% (95% CI, 60% to 98%). Multivariable logistic regression analyses showed that FOXP3 mRNA level remains predictive of TCMR reversal after individual adjustment for serum creatinine measured at time of biopsy, time from transplant to biopsy, and type of organ donor (Table 3).

**Fig. 3.**
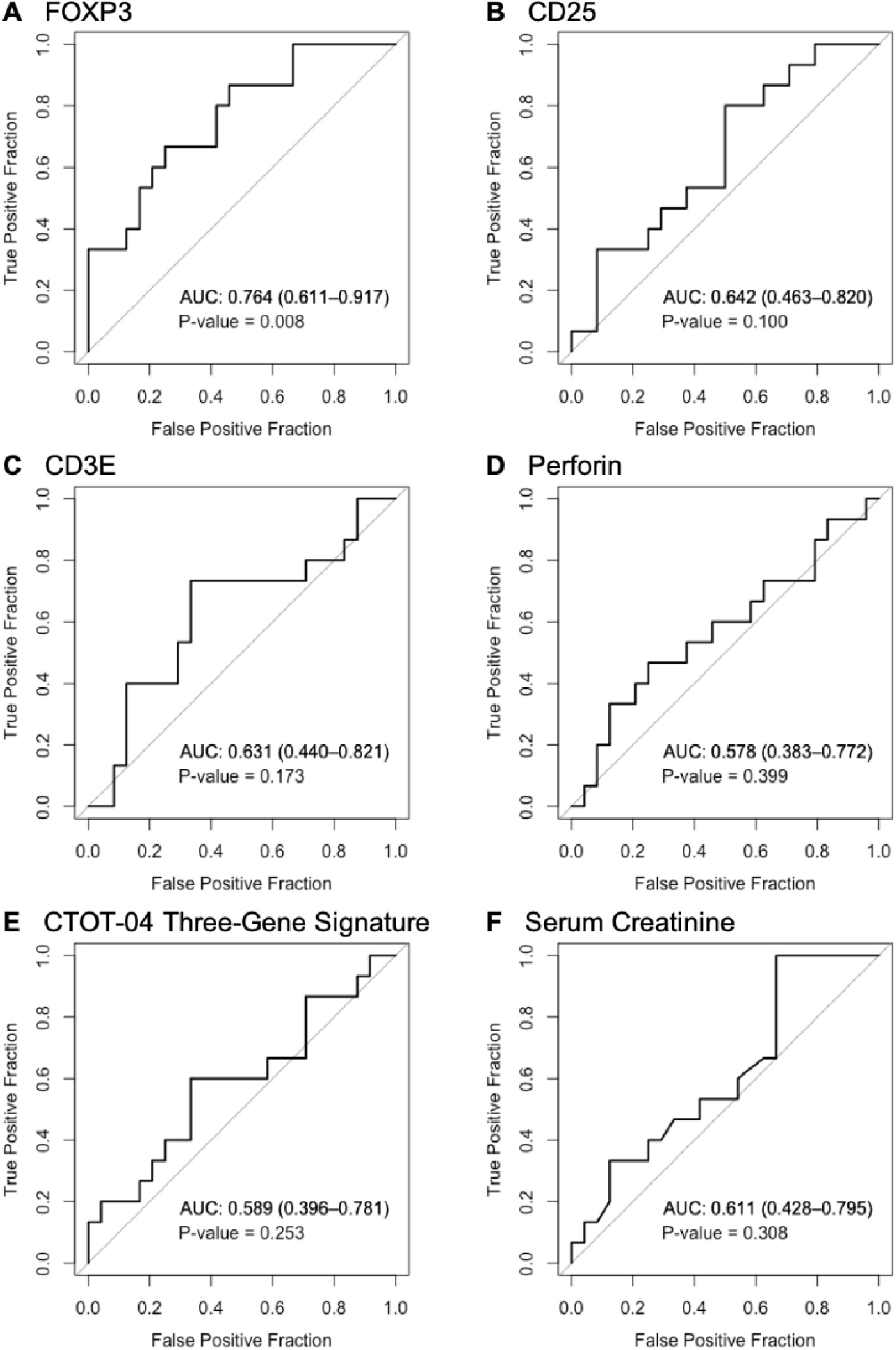
Receiver operating characteristic (ROC) curve analyses. The fraction of true positive results (sensitivity) and the fraction of false positive results (1- specificity) for predicting reversal of an episode of TCMR are shown. (A) The area under the receiver-operating-characteristic curve (AUC) was 0.764 (95% confidence interval [CI], 0.611 to 0.917; *P*=0.008) for 18S ribosomal RNA (rRNA) normalized FOXP3 mRNA. (B) The AUC was 0.642 (95% CI, 0.463 to 0.820; *P*=0.100) for 18S rRNA normalized CD25 mRNA. (C) The AUC was 0.631 (95% CI, 0.440 to 0.821, *P*=0.173) for 18S rRNA normalized CD3E mRNA. (D) The AUC was 0.578 (95% CI, 0.383 to 0.772, *P*=0.399) for 18S rRNA normalized perforin mRNA. (E) The AUC for the CTOT-04 three-gene TCMR diagnostic signature (calculated from 18S rRNA normalized CD3E and IP10 mRNAs and 18S rRNA) was 0.589 (95% CI, 0.396 to 0.781, *P*=0.253). (F) The AUC for serum creatinine level measured at time of TCMR biopsy was 0.611 (95% CI, 0.428 to 0.795, *P*=0.308). AUC is presented in each panel along with corresponding 95% confidence intervals. *P*-values are obtained from Wald tests from logistic regression analyses predicting reversible status from the measure of interest. An AUC value of 0.5 is no better than that expected by chance (the null hypothesis) and a value of 1.0 reflects a perfect discriminator.

**Table 3.**
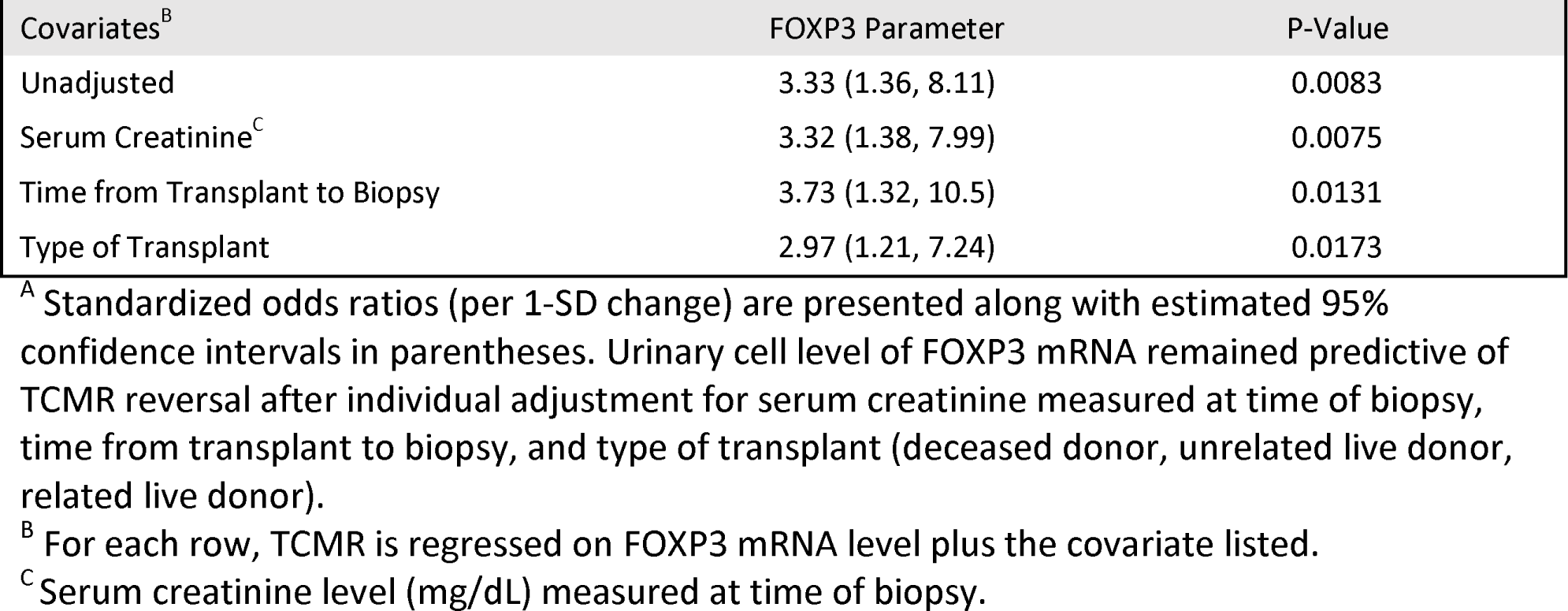
Standardized odds ratios for TCMR reversal: multivariable logistic regression analyses.^A^

|  |  |  |
| --- | --- | --- |
| Covariates<sup>B</sup> | FOXP3 Parameter | P-Value |
| Unadjusted | 3.33 (1.36, 8.11) | 0.0083 |
| Serum Creatinine<sup>C</sup> | 3.32 (1.38, 7.99) | 0.0075 |
| Time from Transplant to Biopsy | 3.73 (1.32, 10.5) | 0.0131 |
| Type of Transplant | 2.97 (1.21, 7.24) | 0.0173 |

<sup>A</sup> Standardized odds ratios (per 1-SD change) are presented along with estimated 95% confidence intervals in parentheses. Urinary cell level of FOXP3 mRNA remained predictive of TCMR reversal after individual adjustment for serum creatinine measured at time of biopsy, time from transplant to biopsy, and type of transplant (deceased donor, unrelated live donor, related live donor).
<sup>B</sup> For each row, TCMR is regressed on FOXP3 mRNA level plus the covariate listed.
<sup>C</sup> Serum creatinine level (mg/dL) measured at time of biopsy.

As previously observed in our single center study (*8*), levels of mRNA for CD25 (AUC=0.642; 95% CI, 0.463 to 0.820; *P*=0.100) (Figure 3B), CD3E (AUC=0.631; 95% CI, 0.440 to 0.821; *P*=0.173) (Figure 3C), and perforin (AUC=0.578; 95% CI, 0.383 to 0.772; *P*=0.399) (Figure 3D) did not predict TCMR reversal in the multicenter CTOT-04 cohort.

The urinary cell three-gene signature of log_10_-transformed 18S rRNA normalized values of CD3E and IP10 mRNAs and log_10_-transformed values of 18S rRNA ([x10^−6^]) was developed and validated in the parent CTOT-04 study to be diagnostic of TCMR (*4*). The three-gene TCMR diagnostic signature did not discriminate reversible from nonreversible TCMR episodes (AUC=0.589; 95% CI, 0.396 to 0.781; *P*=0.253) (Figure 3E). Serum creatinine measured at the time of biopsy did not predict TCMR reversal (AUC=0.611; 95% CI, 0.428 to 0.795; *P*=0.308) (Figure 3F). The use of thymoglobulin as anti-rejection treatment for TCMR (AUC=0.479; 95% CI, 0.321 to 0.637; *P*=0.792) (data not shown) did not discriminate between reversible and nonreversible TCMR.

### Survival analyses of kidney allografts

We examined whether the functional criterion used in this study to classify an episode of TCMR as reversible or nonreversible is associated with kidney allograft survival. Kaplan-Meier survival analysis showed that survival rates of recipients with reversible TCMR are similar to those with No Rejection biopsies, whereas almost all graft losses were in those with nonreversible TCMR biopsies (Figure 4A, *P*<0.0001, Log-rank test).

**Fig. 4.**
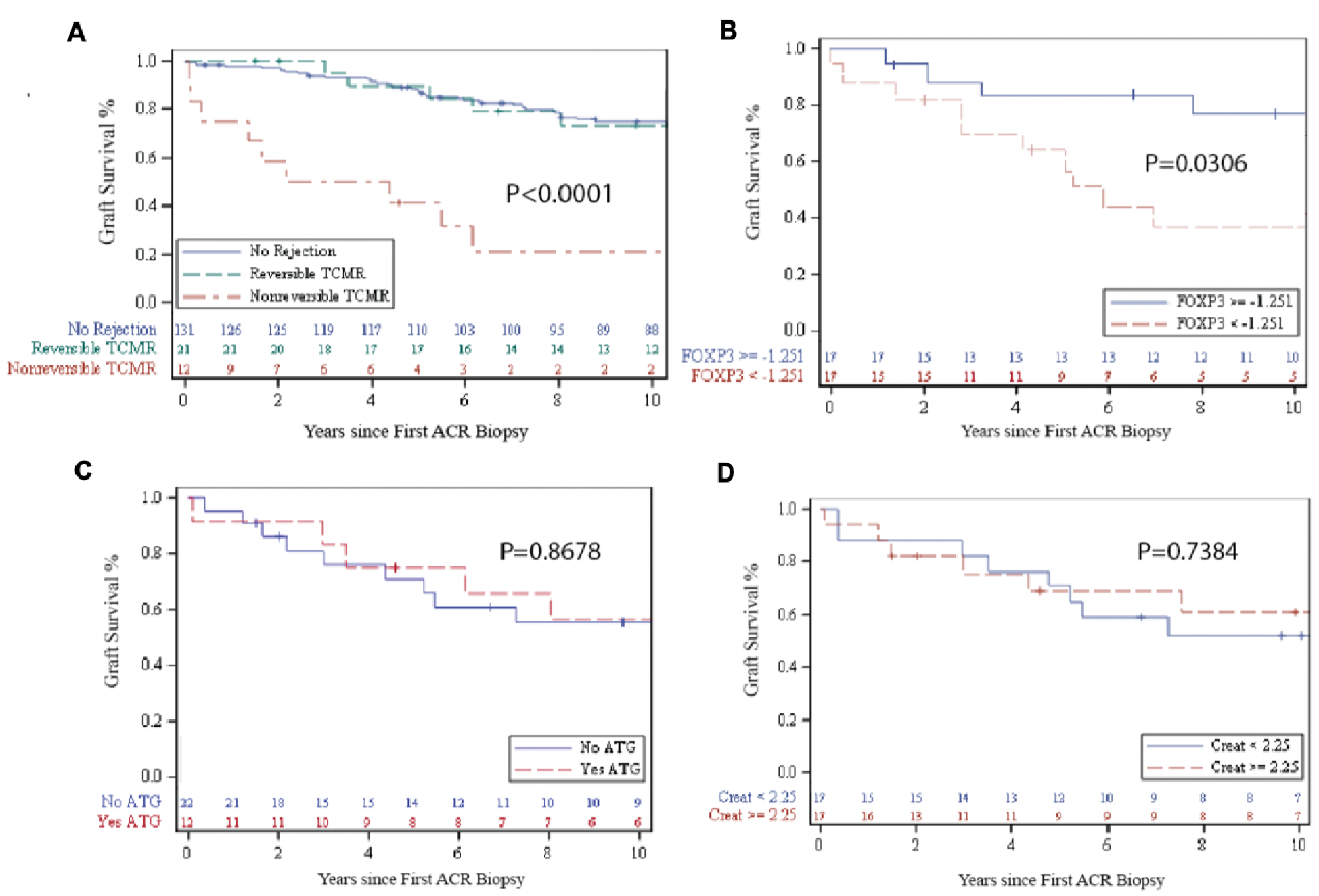
Kaplan-Meier kidney allograft survival curves. (A) Survival curves of patients who had reversible TCMR, nonreversible TCMR, or No Rejection biopsy only. (B) Survival curves for the 34 patients with at least one episode of TCMR, stratified by a median split of log_10_-transformed 18S rRNA normalized FOXP3 mRNA level. (C) Survival curves for the same 34 patients, stratified by anti-rejection therapy with thymoglobulin (Yes ATG) vs. ATG not used for or anti-rejection therapy (No ATG). (D) Survival curves for the same 34 patients, stratified by a median split of serum creatinine measured at time of first TCMR biopsy. Time to event was calculated from date of TCMR biopsy (or the date of last TCMR biopsy in those with multiple episodes) or No Rejection biopsy until graft failure or last follow-up date. Subjects were censored if they experienced death prior to graft failure or were lost to follow-up. *P*-values are derived from log-rank tests. At-risk tables are shown in each plot, just above the X-axis.

Kidney allograft recipients with FOXP3 mRNA levels above the median FOXP3 mRNA level of -1.251 had significantly better graft survival compared to those with levels below the median (Figure 4B, *P*=0.0306). Multivariable Cox proportionate hazards regression analysis showed that FOXP3 mRNA level remains significantly predictive of kidney allograft outcomes after adjustment for serum creatinine measured at time of biopsy, but not after adjustment for time from transplant to biopsy or type of transplant (Table 4).

**Table 4.** Kidney allograft survival: multivariable Cox proportional hazards regression analyses.^A^

|  |  |  |
| --- | --- | --- |
| Covariates<sup>B</sup> | FOXP3 HR | P-Value |
| Unadjusted | 0.55 (0.30, 0.99) | 0.0474 |
| Serum Creatinine<sup>C</sup> | 0.55 (0.30, 0.99) | 0.0475 |
| Time from Transplant to Biopsy | 0.64 (0.30, 1.33) | 0.2309 |
| Type of Transplant | 0.65 (0.36, 1.18) | 0.1584 |

<sup>A</sup> Standardized hazard ratios (per 1-SD change) for kidney allograft survival are presented along with estimated 95% confidence intervals in parentheses. Urinary cell FOXP3 mRNA level remained predictive of graft survival after individual adjustment for serum creatinine measured at time of biopsy but not after adjustment for time from transplant to biopsy, and type of transplant.
<sup>B</sup> For each row, graft survival is regressed on FOXP3 mRNA level plus the covariate listed.
<sup>C</sup> Serum creatinine level (mg/dL) measured at time of biopsy.

Urinary cell levels of mRNA for CD25 (*P*=0.4371), CD3E (*P*=0.8062), and perforin (*P*=0.9839), measured at the time of TCMR biopsy and stratified by their median levels, were not associated with kidney allograft survival. Neither the use of thymoglobulin for anti-rejection therapy (Figure 4C, *P*=0.8678) nor the level of serum creatinine measured at time of biopsy was associated with kidney allograft survival (Figure 4D, *P*=0.7384).

## Discussion

FOXP3+ Tregs play a pivotal role in preventing autoimmunity and maintaining immune homeostasis (*9, 10*). Preclinical studies suggest that Tregs prevent or delay the onset of allograft rejection and may induce tolerance (*11–13*). In our earlier single center clinical study, we found that urinary cell FOXP3 mRNA level is diagnostic of TCMR and predicts TCMR reversal; we also found that urinary cell FOXP3 mRNA level is associated with kidney allograft failure (*8*). In the current investigation, we have replicated these findings using urine samples collected from an external cohort of kidney allograft recipients enrolled in the multicenter CTOT-04 study. Our replication of earlier findings is significant from a biological perspective regarding the potential role of Tregs in regulating kidney allograft rejection, and is reassuring in the context of the existing reproducibility crisis in replicating scientific findings (*16, 17*).

Our profiling identified molecular features consistent with the hypothesis that the balance between T effectors and Tregs is tipped towards Treg deficiency at the onset of nonreversible TCMR. Whereas the levels of mRNA for cytopathic molecule perforin, pan T cell marker CD3E, and T cell activation marker CD25 were not significantly different between urine matched to reversible TCMR biopsies and urine matched to nonreversible TCMR biopsies, level of mRNA for Treg specification factor FOXP3 was significantly lower in urine matched to nonreversible TCMR biopsies compared to urine matched to reversible TCMR biopsies.

Recombinant IL-2 or Tregs have been used to prevent graft versus host disease (GVHD) in recipient hematopoietic stem cell recipients and to prevent acute rejection in solid organ graft rejection (*14, 15, 21*). The findings from the current study suggest that infusion of IL-2 or Tregs may be a consideration for those with TCMR associated with a low urinary cell FOXP3 mRNA. A switch from calcineurin inhibitor (CNI)-based immunosuppression to rapamycin-based immunosuppression could also be considered in this setting given that CNIs are potent inhibitors of IL-2 gene transcription, whereas rapamycin can reprogram Treg metabolism and improve its function (*22–24*).

The overall survival of kidney allografts in the CTOT-04 study cohort was similar to the US kidney graft survival rates (*25*) suggesting that our study participants are representative of the US kidney transplant population. Kidney graft survival was significantly inferior in patients with biopsy-confirmed TCMR, with most graft failures occurring in those with nonreversible TCMR biopsies. Importantly, urinary cell FOXP3 mRNA level, measured at time of TCMR, foretold allograft failure. Laboratory parameters such as serum creatinine, measured at the time of allograft biopsy, or the use T cell depleting agent antithymocyte globulin as anti-rejection therapy did not predict kidney allograft failure in this investigation.

A number of features of our study are worthy of emphasis. First, urinary cell FOXP3 level was assessed in what is likely to be the largest number of urine samples from the largest number of kidney allograft recipients from 5 academic transplant sites. Second, urine samples were collected under the direction of an independent Statistical and Clinical Coordinating Center ensuring protocol-based sample collection, and data entry and integrity (*4*). Third, the oligonucleotide primers we designed for the quantification of mRNAs in the customized RT-QPCR assays obviated DNA amplification and ensured that only mRNAs are measured in the assays. Fourth, our incorporation of preamplification prior to mRNA measurement compensated for the low RNA yield from urinary cells and facilitated successful measurement of multiple mRNAs from the same cDNA sample. Fifth, the use of a customized BAK amplicon in the RT-QPCR assays allowed for absolute quantification of transcript numbers and avoided pitfalls associated with relative quantification (*26*). Finally, our study included a clinically relevant endpoint of graft failure following TCMR.

Our study has a number of shortcomings. We characterized TCMR reversal based on functional recovery rather than by histological confirmation. This may not be a significant limitation since graft outcome in this study was strongly associated with the functional criterion used to classify an episode of TCMR. Another limitation is that we did not assess the functional activity of Tregs and we inferred Treg deficiency based on FOXP3 mRNA abundance. We note that mRNA expression patterns in themselves have helped inform therapeutic decisions (*27*). Our profiling of urinary cells rather than the allograft itself might be considered a limitation as well. This is less of a concern in the light of RNA sequencing demonstrating that kidney allograft biopsy transcriptional signatures are enriched in urinary cells (*28*).

In sum, we have replicated our single center findings that urinary cell FOXP3 mRNA level is diagnostic of TCMR, predicts TCMR reversal and is associated with graft survival following an episode of TCMR. It has been reported that 40% of TCMR episodes fail to respond to state-of-the-art anti-rejection therapy (*5*). Our finding that recalcitrant TCMR is associated with low FOXP3 abundance suggest that infusion of IL-2, a trophic factor for Tregs, infusion of Tregs or rapamycin that may reprogram Treg metabolism may represent novel treatment options for TCMR exemplified by a urinary cell FOXP3 mRNA level consistent with Treg deficiency.

## Data Availability

The data that support the findings of this study are available on request from the corresponding author. The data are not publicly available due to privacy or ethical restrictions.

## Acknowledgments

The authors gratefully acknowledge Dr. Nancy D. Bridges, Transplantation Branch, National Institute of Allergy and Infectious Diseases, Bethesda, MD, USA for her careful review of the manuscript and key insights. The authors thank Ms. Christina Chang and Ms. Christine Hoang (Weill Cornell Medicine, New York, NY) for their superb technical assistance in performing the RT-QPCR assays. The authors thank the United Network for Organ Sharing for providing the clinical data for the calculation of survival curves of kidney allograft recipients. The data reported here have been supplied by UNOS as the contractor for the Organ Procurement and Transplantation Network (OPTN). The OPTN data system includes data on all donors, wait-listed candidates, and transplant recipients in the US, submitted by all members of OPTN. The Health Resources and Services Administration (HRSA), U.S. Department of Health and Human Services provides oversight to the activities of the OPTN contractor. The interpretation and reporting of these data are the responsibility of the authors and in no way should be seen as an official policy of or interpretation by the OPTN or the U.S. Government.

## Author contributions

MS conceived and coordinated the study. DL, PA, JES, and MS wrote the manuscript. DD made critical contributions to the preparation of the manuscript including data collection and kidney allograft survival analysis. RD designed the oligonucleotide primers and TaqMan probes, and developed and supervised the performance of preamplification-enhanced real-time quantitative PCR assays. TM, ML, JL, and FM made significant contributions to data analysis. VKS coordinated data acquisition and management. DL performed the statistical analyses. JES provided critical statistical guidance to DL.

## Supplementary Materials

### Materials and Methods

**Table S1.**
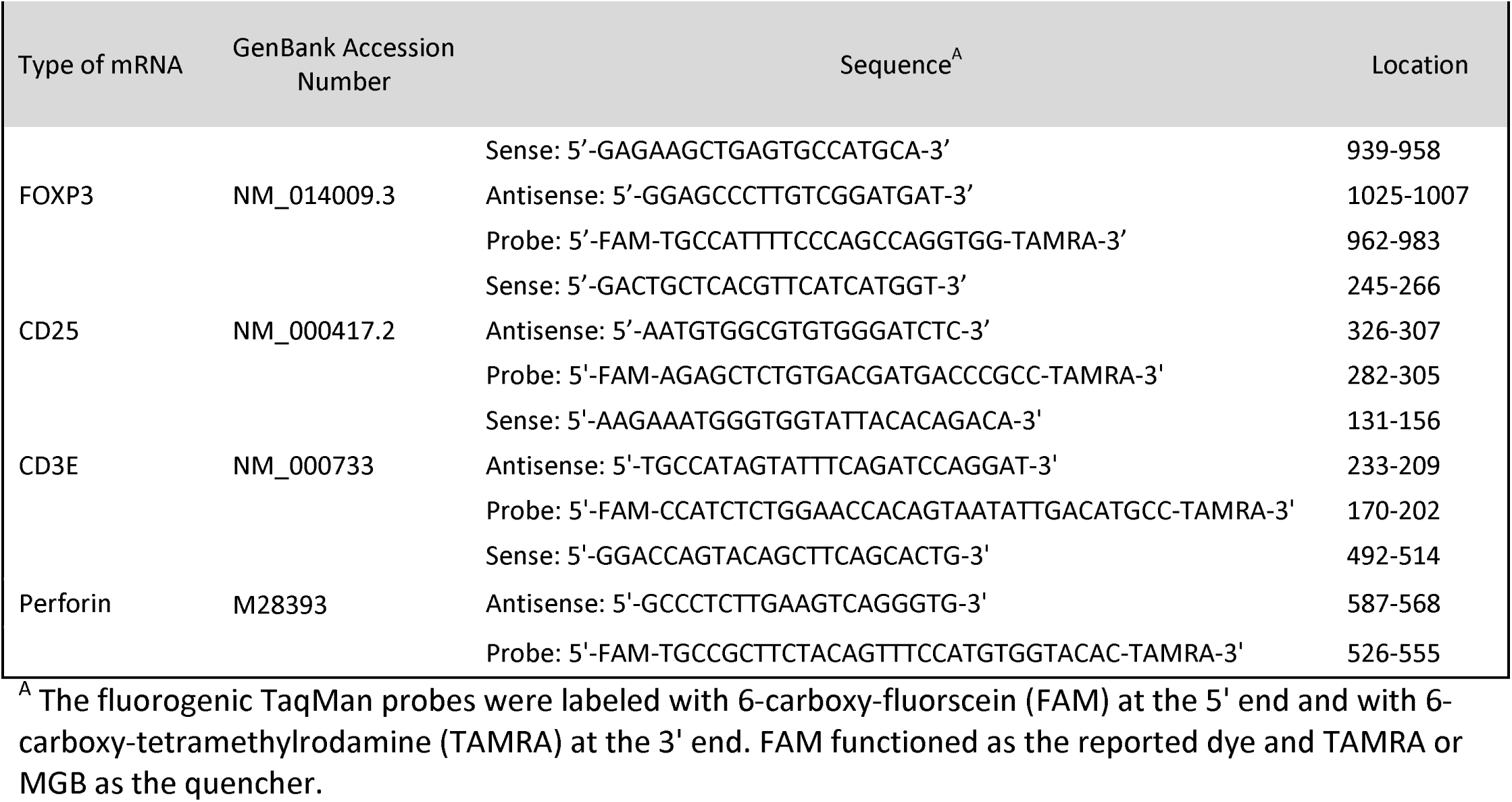
Oligonucleotide primers and TaqMan probes used to measure absolute levels of each mRNA.

**Table S2.**
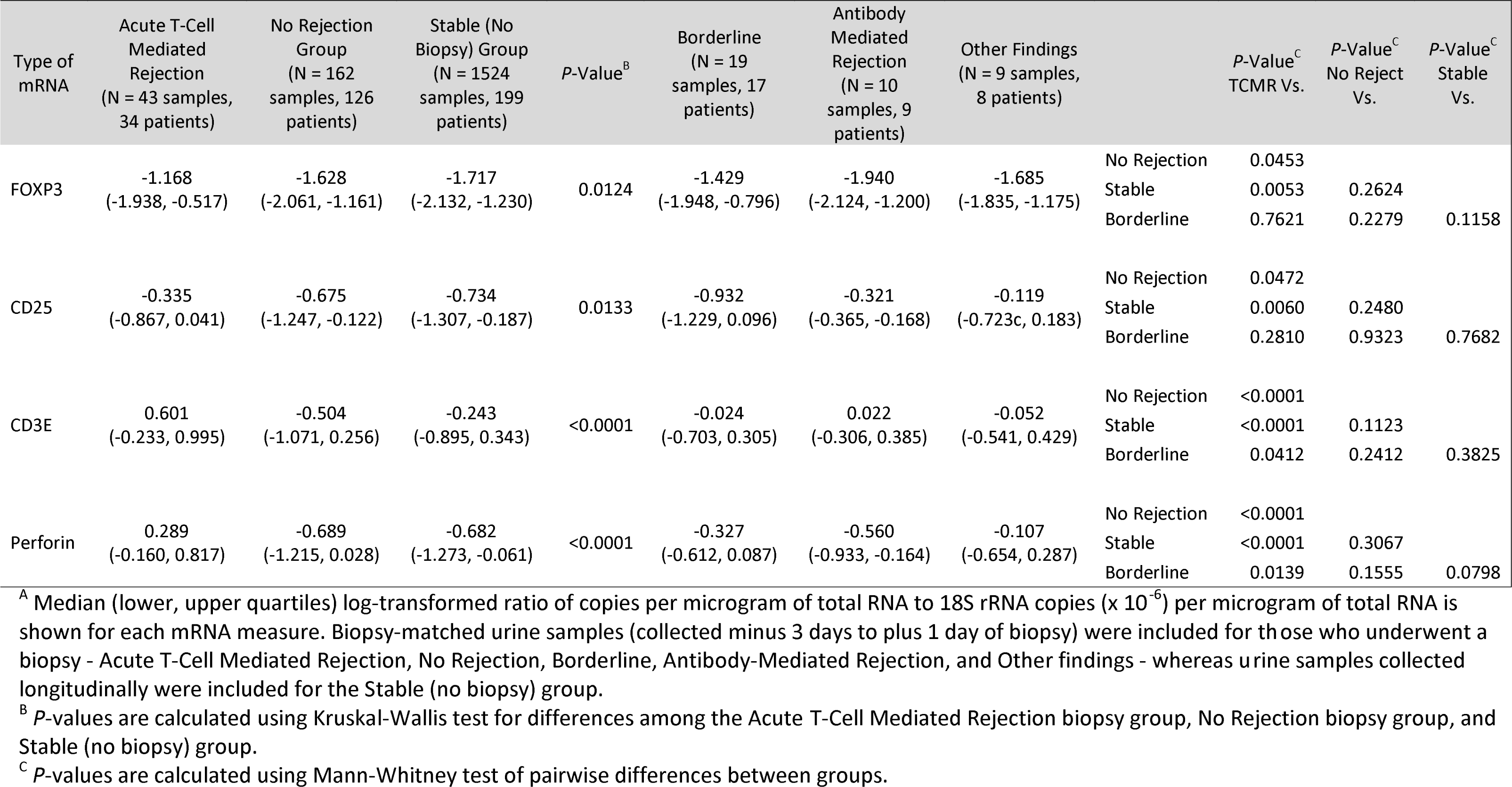
18S rRNA normalized, log_10_-transformed levels of mRNA in urinary cells across diagnostic categories.^A^

**Table S3.** Characteristics of kidney transplant recipients with acute T-cell mediated rejection by reversible status.^A^

| Recipient Characteristics | Total | Reversible TCMR | Nonreversible TCMR |
| --- | --- | --- | --- |
| Study subjects<sup>B</sup> | 33 | 21 | 12 |
| Number of biopsies<sup>C</sup> | 39 | 24 | 15 |
| Age, years |  |  |  |
| Mean (SD) | 45.0 (12.0) | 48.0 (10.9) | 39.8 (12.5) |
| Median | 43 | 46 | 36 |
| Min, Max | 24, 73 | 35, 69 | 24, 73 |
| Gender, N (%) |  |  |  |
| Female | 8 (24.2) | 6 (28.6) | 2 (16.7) |
| Male | 25 (75.8) | 15 (71.4) | 10 (83.3) |
| Ethnicity, N (%) |  |  |  |
| Hispanic or Latino | 2 (6.1) | 1 (4.8) | 1 (8.3) |
| Not Hispanic or Latino | 30 (90.9) | 19 (90.5) | 11 (91.7) |
| Unknown or Not Reported | 1 (3.0) | 1 (4.8) | 0 (0) |
| Race, N (%) |  |  |  |
| Black or African American | 13 (39.4) | 6 (28.6) | 7 (58.3) |
| White | 19 (57.6) | 14 (66.7) | 5 (41.7) |
| Asian | 1 (3.0) | 1 (4.8) | 0 (0) |
| American Indian or Alaska Native | 0 (0) | 0 (0) | 0 (0) |
| Other | 0 (0) | 0 (0) | 0 (0) |
| Unknown or Not Reported | 0 (0) | 0 (0) | 0 (0) |
| Induction Therapy, N (%) |  |  |  |
| IL-2 Receptor Antibody | 6 (18.2) | 5 (23.8) | 1 (8.3) |
| CAMPATH-1H | 9 (27.3) | 7 (33.3) | 2 (16.7) |
| Thymoglobulin | 15 (45.5) | 8 (38.1) | 7 (58.3) |
| More than one induction therapy | 2 (6.1) | 0 (0) | 2 (16.7) |
| No Induction Therapy | 1 (3.0) | 1 (4.8) | 0 (0) |
| Missing Information | 0 (0) | 0 (0) | 0 (0) |
| BMI |  |  |  |
| Mean (SD) | 30.6 (6.3) | 30.8 (6.7) | 30.2 (5.6) |
| Median | 30 | 29 | 30 |
| Min, Max | 22, 43 | 22, 43 | 24, 38 |
| < 18.5 | 0 (0) | 0 (0) | 0 (0) |
| 18.5 - 24.9 | 9 (27.3) | 6 (28.6) | 3 (25.0) |
| 25.0 - 29.9 | 6 (18.2) | 4 (19.1) | 2 (16.7) |
| ≥ 30.0 | 14 (42.4) | 10 (47.6) | 4 (33.3) |
| Missing | 4 (12.1) | 1 (4.8) | 3 (25.0) |
| Donor Characteristics |  |  |  |
| Age, years |  |  |  |
| Mean (SD) | 41.8 (11.7) | 40.6 (12.4) | 44.0 (10.5) |
| Median | 40 | 40 | 43 |
| Min, Max | 20, 65 | 20, 65 | 26, 61 |
| Missing | 0 | 0 | 0 |
| Gender, N (%) |  |  |  |
| Female | 17 (51.5) | 10 (47.6) | 7 (58.3) |
| Male | 16 (48.5) | 11 (52.4) | 5 (41.7) |
| Ethnicity, N (%) |  |  |  |
| Hispanic or Latino | 3 (9.1) | 2 (9.5) | 1 (8.3) |
| Not Hispanic or Latino | 29 (87.9) | 19 (90.5) | 10 (83.3) |
| Unknown or Not Reported | 1 (3.0) | 0 (0) | 1 (8.3) |
| Race, N (%) |  |  |  |
| Black or African American | 10 (30.3) | 7 (33.3) | 3 (25.0) |
| White | 21 (63.6) | 13 (61.9) | 8 (66.7) |
| Asian | 1 (3.0) | 0 (0) | 1 (8.3) |
| American Indian or Alaska Native | 0 | 0 (0) | 0 (0) |
| Other | 0 | 0 (0) | 0 (0) |
| Unknown or Not Reported | 1 (3.0) | 1 (4.8) | 0 (0) |
| Source of Donor, N (%) |  |  |  |
| Deceased | 15 (45.5) | 6 (28.6) | 9 (75.0) |
| Living/related | 8 (24.2) | 8 (38.1) | 0 (0) |
| Living/unrelated | 10 (30.3) | 7 (33.3) | 3 (25.0) |
| Cause of Death, N (%) |  |  |  |
| Anoxia | 4 (26.7) | 2 (9.5) | 2 (15.4) |
| Cerebrovascular Accident/Injury/Stroke | 7 (46.7) | 2 (9.5) | 5 (38.5) |
| Head Injury/Trauma | 2 (13.3) | 1 (4.8) | 1 (7.7) |
| Intracranial Bleed | 1 (6.7) | 0 (0) | 1 (7.7) |
| Motor Vehicle Accident | 0 (0) | 0 (0) | 0 (0) |
| Other | 0 (0) | 0 (0) | 0 (0) |
| Unknown | 0 (0) | 0 (0) | 0 (0) |
| Missing | 1 (6.7) | 1 (4.8) | 0 (0) |

<sup>A</sup> An episode of TCMR was classified as reversible if the serum creatinine level returned to within 15% of pre-rejection levels within 4 weeks after initiation of anti-rejection therapy.
<sup>B</sup> Thirty-three subjects contributed 39 TCMR biopsy matched urine samples. The subjects were classified based on their last TCMR biopsy episode being reversible or nonreversible.
<sup>C</sup> Three patients had multiple biopsies with each contributing three biopsy-matched urine samples. One of the 3 patients contributed one urine sample matched to a nonreversible episode based on absence of improvement in serum creatinine followed by two urine samples matched to two episodes of reversible TCMR. The two remaining patients contributed one urine sample matched to a reversible episode of TCMR followed by two urine samples matched to two episodes of nonreversible TCMR each (five months apart in one patient and 12 months apart in the second patient).

**Table S4.** Biopsy associated characteristics of kidney transplant recipients with acute T-cell mediated rejection by reversible status.^A^

|  | Total | Reversible TCMR | Nonreversible TCMR |
| --- | --- | --- | --- |
| Study subjects<sup>B</sup> | 33 | 21 | 12 |
| Number of biopsies<sup>C</sup> | 39 | 24 | 15 |
| Time from Transplant to Biopsy, days |  |  |  |
| Mean (SD) | 217 (188) | 159 (160) | 310 (196) |
| Median | 180 | 117 | 269 |
| Min, Max | 3, 701 | 3, 491 | 18, 701 |
| Serum Creatinine at Baseline, mg/dL |  |  |  |
| Mean (SD) | 1.9 (1.2) | 2.0 (1.4) | 1.7 (0.5) |
| Median | 1.6 | 1.6 | 1.6 |
| Min, Max | 0.9, 8.2 | 0.9, 8.2 | 1.1 (3.3) |
| Serum Creatinine at Time of Biopsy, mg/dL |  |  |  |
| Mean (SD) | 3.1 (2.8) | 2.8 (2.3) | 3.7 (3.4) |
| Median | 2.3 | 2.3 | 2.5 |
| Min, Max | 1.1, 13.3 | 1.1, 12.2 | 1.7, 13.3 |
| Serum Creatinine after 4 weeks, mg/dL |  |  |  |
| Mean (SD) | 2.2 (1.2) | 1.8 (1.2) | 2.8 (0.9) |
| Median | 1.9 | 1.6 | 2.7 |
| Min, Max | 0.9, 6.6 | 0.9, 6.6 | 1.6, 4.5 |
| Anti-Rejection Regimen<sup>D</sup> |  |  |  |
| Glucocorticoids | 37 | 23 | 14 |
| Antilymphocyte antibodies | 14 | 9 | 5 |
| Other | 6 | 2 | 4 |

<sup>A</sup> An episode of TCMR was classified as reversible if the serum creatinine level returned to within 15% of pre-rejection levels within 4 weeks after initiation of anti-rejection therapy.
<sup>B</sup> Thirty-three subjects contributed 39 TCMR biopsy matched urine samples. The subjects were classified based on their last TCMR biopsy episode being reversible or nonreversible.
<sup>C</sup> Three patients had multiple biopsies with each contributing three biopsy matched urine samples. One of the 3 patients contributed one urine sample matched to a nonreversible episode based on absence of improvement in serum creatinine followed by two urine samples matched to two episodes of reversible TCMR. The two remaining patients contributed one urine sample matched to a reversible episode of TCMR followed by two urine samples matched to two episodes of nonreversible TCMR each (five months apart in one patient and 12 months apart in the second patient).
<sup>D</sup> The sum of anti-rejection treatments for biopsies within a particular column is not equal to the total number of biopsies (TCMR diagnoses) because several TCMR episodes were treated with multiple anti-rejection regimens.

## References

1. G. Opelz, B. Dohler, R. Collaborative Transplant Study, Influence of time of rejection on long-term graft survival in renal transplantation. Transplantation 85, 661–666 (2008).

2. M. El Ters et al, Kidney allograft survival after acute rejection, the value of follow-up biopsies. Am J Transplant 13, 2334–2341 (2013).

3. D. E. Hricik et al, Multicenter validation of urinary CXCL9 as a risk-stratifying biomarker for kidney transplant injury. Am J Transplant 13, 2634–2644 (2013).

4. M. Suthanthiran et al, Urinary-cell mRNA profile and acute cellular rejection in kidney allografts. N Engl J Med 369, 20–31 (2013).

5. Y. Bouatou et al., Response to treatment and long-term outcomes in kidney transplant recipients with acute T cell-mediated rejection. Am J Transplant, (2019).

6. P. N. Furness, N. Taub, P. Convergence of European Renal Transplant Pathology Assessment Procedures, International variation in the interpretation of renal transplant biopsies: report of the CERTPAP Project. Kidney Int 60, 1998–2012 (2001).

7. P. N. Furness et al, Protocol biopsy of the stable renal transplant: a multicenter study of methods and complication rates. Transplantation 76, 969–973 (2003).

8. T. Muthukumar et al, Messenger RNA for FOXP3 in the urine of renal-allograft recipients. N Engl J Med 353, 2342–2351 (2005).

9. S. Sakaguchi et al., Regulatory T Cells and Human Disease. Annu Rev Immunol, (2020).

10. P. Georgiev, L. M. Charbonnier, T. A. Chatila, Regulatory T Cells: the Many Faces of Foxp3. J Clin Immunol 39, 623–640 (2019).

11. S. P. Cobbold et al, Induction of foxP3+ regulatory T cells in the periphery of T cell receptor transgenic mice tolerized to transplants. J Immunol 172, 6003–6010 (2004).

12. M. H. Albert, Y. Liu, C. Anasetti, X. Z. Yu, Antigen-dependent suppression of alloresponses by Foxp3-induced regulatory T cells in transplantation. Eur J Immunol 35, 2598–2607 (2005).

13. W. Li et al, Bronchus-associated lymphoid tissue-resident Foxp3+ T lymphocytes prevent antibody-mediated lung rejection. J Clin Invest 129, 556–568 (2019).

14. M. Romano, G. Fanelli, C. J. Albany, G. Giganti, G. Lombardi, Past, Present, and Future of Regulatory T Cell Therapy in Transplantation and Autoimmunity. Front Immunol 10, 43 (2019).

15. L. M. R. Ferreira, Y. D. Muller, J. A. Bluestone, Q. Tang, Next-generation regulatory T cell therapy. Nat Rev Drug Discov 18, 749–769 (2019).

16. M. Baker, Is there a reproducibility crisis? Nature 533, 452–454 (2016).

17. M. McNutt, Reproducibility. Science 343, 229 (2014).

18. C. T. Le, A solution for the most basic optimization problem associated with an ROC curve. Stat Methods Med Res 15, 571–584 (2006).

19. X. Robin et al, pROC: an open-source package for R and S+ to analyze and compare ROC curves. BMC Bioinformatics 12, 77 (2011).

20. L. C. Racusen et al, Antibody-mediated rejection criteria - an addition to the Banff 97 classification of renal allograft rejection. Am J Transplant 3, 708–714 (2003).

21. J. S. Whangbo, J. H. Antin, J. Koreth, The role of regulatory T cells in graft-versus-host disease management. Expert Rev Hematol 13, 141–154 (2020).

22. J. Koreth et al, Interleukin-2 and regulatory T cells in graft-versus-host disease. N Engl J Med 365, 2055–2066 (2011).

23. A. A. Kennedy-Nasser et al, Ultra low-dose IL-2 for GVHD prophylaxis after allogeneic hematopoietic stem cell transplantation mediates expansion of regulatory T cells without diminishing antiviral and antileukemic activity. Clin Cancer Res 20, 2215–2225 (2014).

24. N. M. Chapman, H. Chi, mTOR signaling, Tregs and immune modulation. Immunotherapy 6, 1295–1311 (2014).

25. A. Hart et al, OPTN/SRTR 2017 Annual Data Report: Kidney. Am J Transplant 19 Suppl 2, 19–123 (2019).

26. T. D. Schmittgen, K. J. Livak, Analyzing real-time PCR data by the comparative C(T) method. Nat Protoc 3, 1101–1108 (2008).

27. S. Paik et al, A multigene assay to predict recurrence of tamoxifen-treated, node-negative breast cancer. N Engl J Med 351, 2817–2826 (2004).

28. A. Verma, T. Muthukumar, Y. Yang, M. Suthanthiran, Urinary cell transcriptomics and acute rejection in human kidney allografts. JCI Insight, (In Press).

